# Reductions in malaria cases after deployment of dual-active ingredient insecticide treated nets (ITNs) in Ghana – a Bayesian interrupted time series analysis

**DOI:** 10.1101/2025.08.20.25334128

**Authors:** Samuel Kweku Oppong, Otubea Owusu-Akrofi, Christian Atta-Obeng, Sylvester Coleman, Punam Amratia, Tasmin L Symons, Kefyalew Addis Alene, Nana Yaw Peprah, Peter W. Gething, Keziah Lawrencia Malm

## Abstract

**Background:** Insecticide treated nets (ITNs) represent a key tool in reducing human vector contact for malaria control. However, increasing insecticide resistance of malaria vectors threatens the effectiveness of pyrethroid-only nets in reducing malaria risk. Next-generation nets, such as those with dual active ingredients, have been recommended for use in areas with high malaria burden and confirmed pyrethroid resistance. Here, we assessed the impact of the distribution of Interceptor^®^ G2 (IG2) ITNs on malaria cases in the Western North region of Ghana distributed in 2021.

**Methods:** We analysed monthly numbers of confirmed malaria cases reported by health facilities in the Western North Region from 2018 to 2023. To control for possible confounding effects of climatic conditions, monthly mean values of both modelled vector habitat suitability and temperature suitability for the periods were included. Bayesian Poisson regression time series models were developed to assess the immediate and sustained impact of IG2 ITNs on malaria case trends.

**Results:** Malaria cases reduced by 30% [odds ratio: 0.70, 95% CrI (0.624, 0.778)] immediately after the distribution of IG2 ITNs in the Western North region. This effect was sustained at six months up to 30 months post-intervention, where cases were reduced by 26% [odds ratio: 0.74, (0.65, 0.75) and 40% [odds ratio: 0.598 (0.495, 0.722)], respectively. The intervention was also strongly associated with reductions in malaria cases in seven of the nine districts in the region, after controlling for climatic factors.

**Conclusion:** This study demonstrates the effectiveness of dual active Interceptor^®^ G2 ITNs in the Western North region, an area with confirmed pyrethroid resistance. These findings support the scale-up of these next-generation nets by National Malaria Programs and highlight the need for further research to explore the utility of these nets in other high-burden malaria areas with region-specific insecticide resistance profiles.

**Key messages:** Malaria prevalence and incidence both decreased after deployment of IG2 ITNs in the Western North region of Ghana.

Distribution of IG2 ITNs caused immediate and sustained impact on malaria case reduction.

Dual-active ingredient insecticide-treated nets are effective in field settings and could be deployed at a large-scale.

## Introduction

Globally, malaria continues to pose a major public health threat as one of the leading causes of illness and death, especially in sub-Saharan Africa(1). Interrupting mosquito-human contact through vector control is a key strategy in malaria control and elimination(2). Insecticide treated nets (ITNs) and indoor residual spraying (IRS) are two core vector control interventions recommended by the World Health Organization (WHO) to prevent malaria transmission(1). The use of ITNs in particular, has been shown to reduce all-cause child mortality, incidences of uncomplicated *Plasmodium falciparum* malaria, and *P. falciparum* malaria prevalence, when compared with untreated nets or no nets(3). Pyrethroids are the preferred insecticides for ITNs (3,4) but widespread resistance in malaria vectors threatens the efficacy of ITNs(2). To mitigate this challenge, next-generation nets treated with piperonyl butoxide (PBO) combined with pyrethroids, as well as dual-active ingredient (dual AI) nets (pyrethroid-pyriproxyfen and pyrethroid-chlorfenapyr) have been introduced, and these have all been shown to boost the efficacy of ITNs against resistant malaria vectors(5). Dual AI nets have shown significant effect in reducing malaria prevalence and clinical episodes, compared with standard pyrethroid-only ITNs(6). It is estimated that 13 million malaria cases and 24,600 deaths have been averted by the introduction of dual AI nets in 17 malaria endemic countries between 2019 and 2022(7).

Pyrethroid-only ITNs have been distributed in Ghana since 2003(8). Upon the adoption of a universal coverage of intervention policy in 2010, four nationwide ITN distribution campaigns have been carried out between 2010 and 2021 aimed at making nets accessible to all populations at risk of malaria(9). In the 2021 campaign, a mixture of pyrethroid-only, pyrethroid+PBO and dual AI nets were distributed, based on recommendations from a subnational stratification exercise conducted in 2019 by WHO(10). The Western North Region, which was part of the Western Region in 2019, had the highest malaria prevalence out of the 10 regions of the country(11) was assigned dual active Interceptor^®^ G2 nets (IG2) AI nets in the 2021 mass ITN distribution(10). Remarkable reductions in malaria prevalence and cases were documented in 2022, suggesting possible reductions in malaria burden post-intervention(12).

Guided by such insight, we hypothesized that the distribution of IG2 nets contributed to a decrease in malaria cases in the Western North Region. To address this, we evaluated the impact of IG2 net distribution on clinical malaria cases in the Western North Region of Ghana from 2018 to 2023 by 1) assessing the magnitude of decrease in clinical malaria cases immediately after the ITN distribution in the region, and 2) assessing the sustained effect up to 30 months after the intervention.

## Methods

### Study region

The Western North Region of Ghana was created out of the Western Region and inaugurated in 2019(13–15). The Region is bordered to the North by the Ashanti and Ahafo Regions, the existing Western Region to the south, as well as by the Central and Ashanti Regions to the East and to the West by the Ivory Coast(14). The Region is divided into nine (9) administrative districts, as shown (Figure 1). The total land size of the Region is estimated as 10 257 km^2^, with a population of 880 921 that represents 2.9% of the national population, according to the 2021 population and housing census(15). The major economic activities in the region are cocoa farming and gold mining, with numerous small- and large-scale mining companies. The Region has about 75% of its vegetation within the high forest zone of Ghana with moderate temperatures ranging from 22°C at night-time and 34°C at daytime. The highest recorded annual rainfall periods in Ghana occur in two peaks, in May-to-July and in September-to-October(14,16). However, intermittent rainfall is noted all year-round, which contributes to a setting that is conducive for perennial transmission of malaria. The Western North Region, which was part of the Western Region in 2019 had a malaria parasite prevalence rate of 27% and this reduced to 4% in 2022(11,12). The main malaria control interventions in the Region are case management, distribution of ITNs through routine channels and periodic mass ITN campaigns(17). Insecticide resistance data from 2015 shows high pyrethroid resistance intensity to malaria vector populations in the Region. Table 1 highlights changes in some malaria indicators before and after the distribution of IG2 ITNs in the Region in 2021.

**Figure 1:**
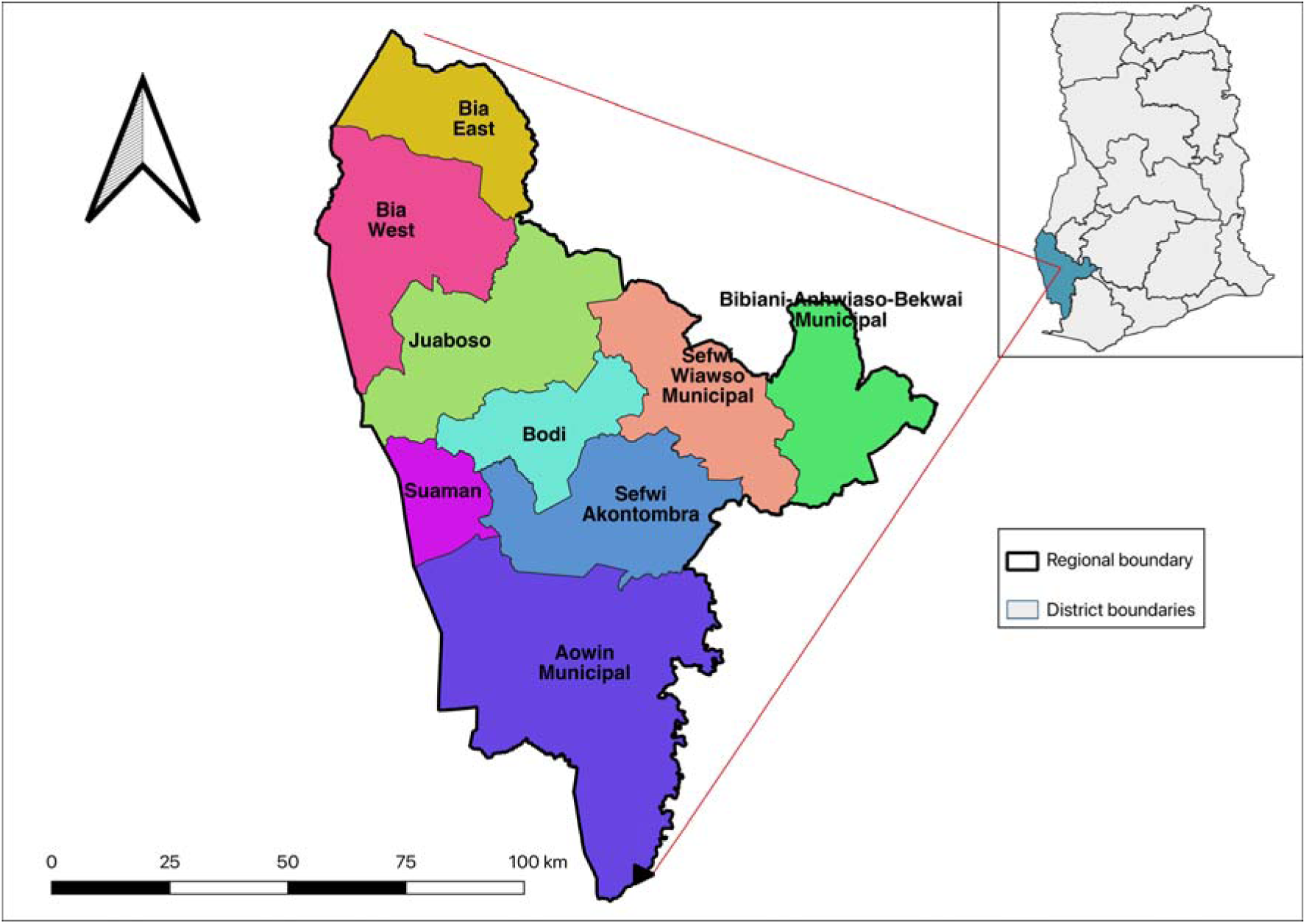
Map of the Western North Region of Ghana showing the locations of all nine Districts. Insert - Map of Ghana showing location of Western North Region (blue).

**Figure 2:**
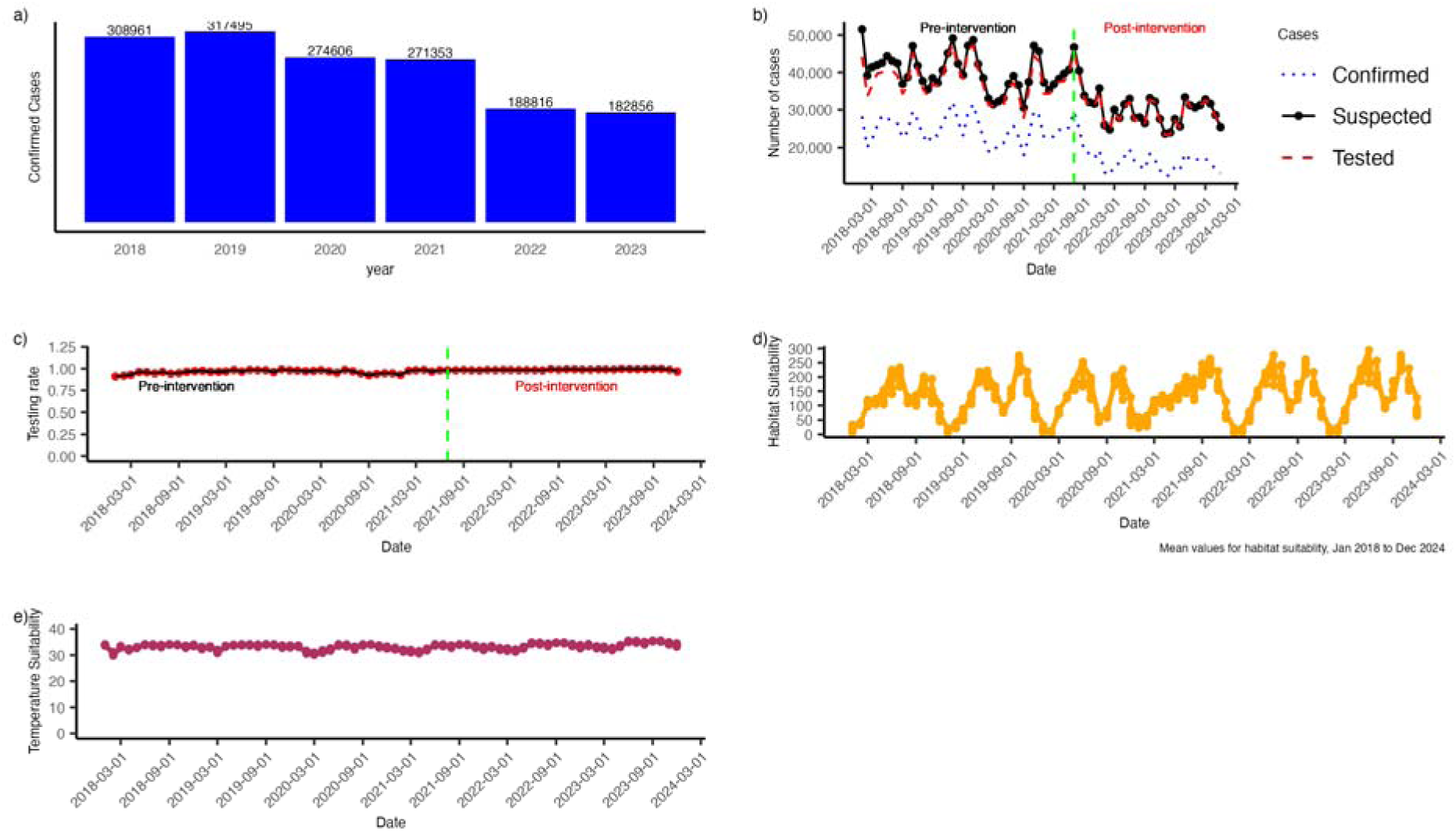
Descriptive analysis of the data. a) Yearly trend of confirmed cases in Western North region, 2018 - 2023. The figure labels on top of the bars represent the total number of cases reported for that year. b) Trend of suspected, tested and confirmed malaria cases in the Western north region, January 2018 to December 2023. Solid black line shows suspected malaria cases, dashed red lines shows cases tested parasitologically and blue dotted dots shows monthly confirmed cases from January 2018 to December 2023. c) Trend of testing rate in the Western north region, January 2018 to December 2023. The green vertical line shows the month the intervention was implemented. d) Trend of habitat suitability (orange lines) and e) - Temperature suitability (blue line) from January 2018 to December 2024.

**Table 1:**
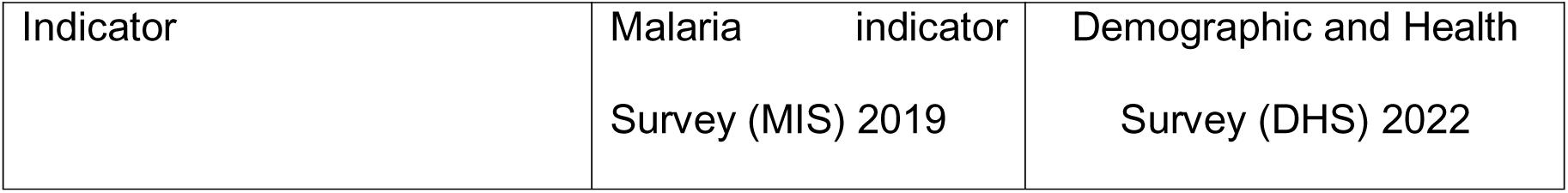
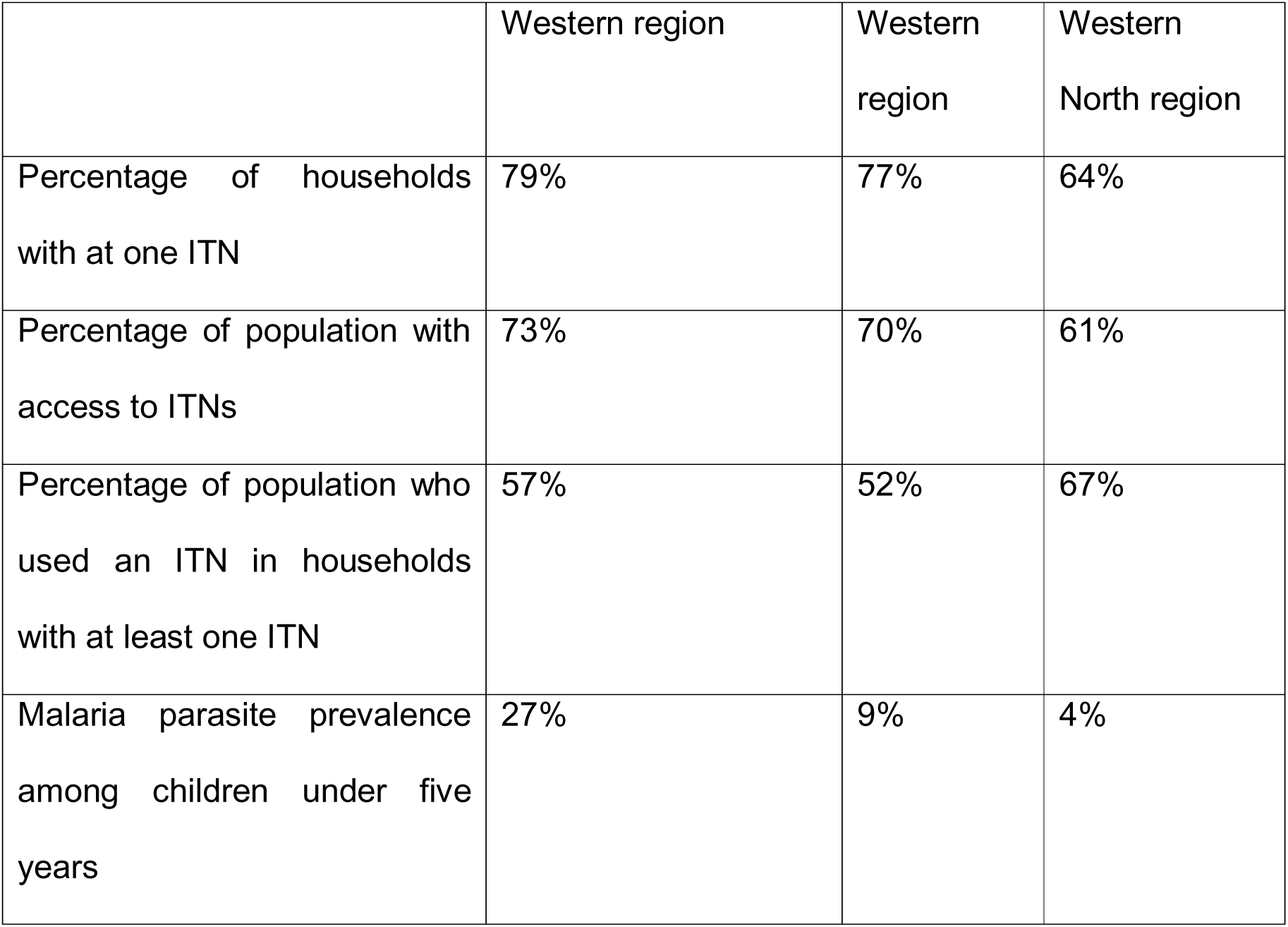
Results from 2019 Malaria Indicator Survey and 2022 Demographic and Health Survey for Western and Western North regions. The Western Region was split into Western and Western North regions after the 2019 MIS.

| Indicator | Malaria indicator<br>Survey (MIS) 2019 | Demographic and Health<br>Survey (DHS) 2022 |
| --- | --- | --- |

|  | Western region | Western region | Western North region |
| --- | --- | --- | --- |
| Percentage of households with at one ITN | 79% | 77% | 64% |
| Percentage of population with access to ITNs | 73% | 70% | 61% |
| Percentage of population who used an ITN in households with at least one ITN | 57% | 52% | 67% |
| Malaria parasite prevalence among children under five years | 27% | 9% | 4% |

### Data

We extracted aggregated malaria data from the District Health Information Management System (DHIMS-2) of the Ghana Ministry of Health(18). DHIMS2 is the primary data reporting platform for health facilities rendering clinical and public health services in the country. The platform is password-protected with access granted to authorized data entry staff and health managers. Malaria and other clinical services rendered in health facilities are either recorded in standardized paper-based registers or electronic medical records systems. Malaria-related data generated and reported by health facilities include number of suspected malaria cases (suspected), number of suspected malaria cases tested for parasites (tested) – either by microscopy or malaria rapid diagnostic test (mRDT), number of tested cases positive for malaria parasites (confirmed) and number of malaria cases treated not tested but treated (presumed). Reported data in DHIMS2 are verified and validated by various levels at specific timepoints in the ensuing month after which the data is ready to be analysed for decision-making.

For this study, we extracted data on suspected, tested and confirmed malaria cases from January 2018 to December 2023 for all the nine districts in the Region. The primary outcome variable used in the analysis was the number of confirmed malaria cases.

### Intervention – Mass ITN distribution campaign

The “intervention” in this study refers to the deployment of dual AI ITNs (that is, IG2 ITNs) in the Western North region. In July 2021, approximately 700 000 interceptor G2 (IG2) ITNs, were distributed to approximately 260 000 households in the Western North Region (see SI-methods for more information on the distribution). In the following sections of this paper, we use the term “dual AI” to represent “IG2 nets”.

### Confounding/potential bias variables

We considered habitat suitability – a proxy for rainfall and temperature suitability as confounding climatic factors that affect malaria in the region. The Habitat Suitability Index (HSI) assesses the ability of a natural habitat to support the development of an organism(19). This covariate incorporates the availability of transient water bodies that facilitates the breeding of mosquitoes and accounts for the aquatic life cycle of the mosquito from laying of the eggs to the emergence of the adult mosquito. It also takes into consideration lag rainfall and its relationship to the development of the mosquito. A Temperature Suitability Index (TSI) is a measure that quantify the impact of air temperature on mosquito survival and development of sporozoite in the mosquito after it bites an infected person(20). It depicts the conduciveness of the temperature in a particular environment to the development and lifespan of adult mosquitoes. TSI relies on the temperature dependent mechanisms in the malaria transmission cycle and combines it with temporal resolution temperature data in its processing(21). We extracted the climatic covariates at 5km resolution for all nine districts in the Region from January 2018 to December 2023. The two climatic covariates were standardized using the scale() function in R(22) prior to model fitting.

### Exploratory analysis

We reviewed the extracted surveillance data for missingness and consistency. We plotted line graphs of suspected, tested and confirmed cases to visualize and identify if any of the following scenarios were present: tested cases greater than (>) suspected cases; confirmed cases > tested cases or suspected cases; treated cases > suspected cases. We also visualised monthly testing rates to examine whether there had been any systematic changes in the testing of suspected cases over the period. Testing rate was defined as:

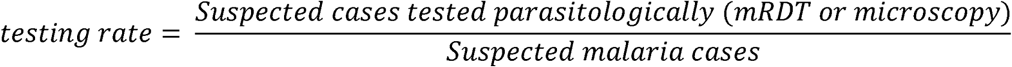

### Bayesian time series model

We used the R-INLA package to build Bayesian interrupted time series (BITS) models, with monthly confirmed malaria cases as the outcome. We built two models to enable us to assess the immediate effect of the intervention differently from the sustained effect of the intervention. In the first model, we included time trend, HSI and TSI as regression terms. The relationship between the exposure (treatment) and outcome after the intervention was modelled as a “step change” impact: an indicator coded as zero before the exposure and one afterwards. Time and climatic covariates were modelled as linear trends over time. We expected a plateau in numbers of malaria cases before the intervention and a sudden decrease immediately afterwards.

In the second BITS model, monthly count of confirmed malaria cases was the outcome, with time trend, time-since-treatment – a six-monthly multi-factor variable for months with five levels, HSI and TSI as explanatory time series. This was to evaluate the magnitude of impact at each time-point independently. We modelled the changing relationship between the outcome at six-monthly time points after the intervention as ‘slope change’ impact. Each time-point was coded as zero at the time of intervention or after that time-point, and one at that time point (See SI- BITS Model for details).

In both models, we included month as a random effect and modelled it as independent and identically distributed (iid), with some specified hyperparameters. We chose a penalized complex prior for the model to control for overfitting (see methods in SI). In the following Results section below, we explore the change in cases due to the intervention by presenting the posterior means and 95% Credible Interval (CrI) attributes for our data.

## Results

### Descriptive analysis

The total number of confirmed cases reported by the region ranged from 308 961 in 2018 to 182 856 in 2023. Confirmed cases reduced slightly from 317 495 in 2019 to

274 606 in 2020 but drastically from 271 353 in 2021 to 188 816 in 2022 (Figure 1a). There was a remarkable reduction in the monthly number of suspected malaria cases and number of confirmed cases after the intervention, with almost all suspected cases receiving a parasitological test (Figure 1b). Testing rates increased from 90% in January 2018 to almost 100% during the periods dual AI nets were deployed (Figure 1c). The time trends of habitat suitability and temperature suitability are also shown in Figures 1d and 1e. There were profound variations in monthly estimates of habitat suitability while estimates for temperature suitability were stable over time.

Bibiani-Anhwiaso-Bekwai reported most of the cases in the Region, with Bia East reporting the least number of cases (SI Figure 1). In all districts, there was a reduction in cases in 2022 compared to 2021. The monthly trend of cases shows a reduction after the intervention in all but one (Suaman) of the 9 districts in the region (Figure 3). The highest declines were observed in Juaboso, Bia West, Sefwi-Wiawso, and Bibiani-Anhwiaso-Bekwai districts.

**Figure 3:**
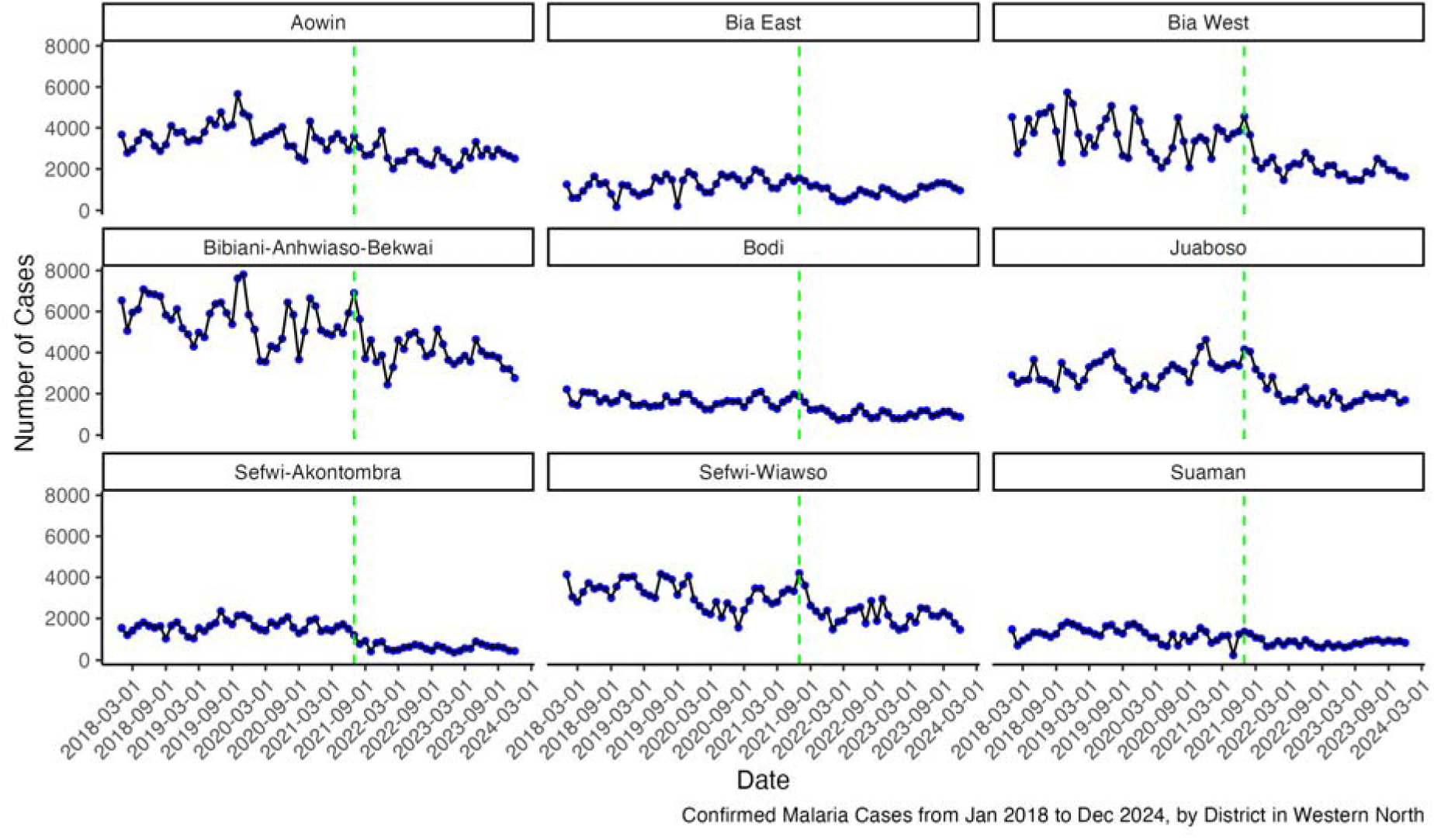
Trends for confirmed malaria cases for all 9 Districts in the Western North Region. Black solid lines with blue dots show the monthly number of cases while the green vertical lines denote the timepoint when the intervention was implemented.

### Model results

The result of the first Bayesian time series model is shown in Table 1. The baseline number of confirmed cases (intercept) before the intervention was 26 991. The intervention was associated with an immediate reduction of confirmed malaria cases by 30% (OR – 0.70, CrI: 0.62, 0.78). The time parameter which signifies the underlining trend or pattern of cases before the intervention showed no influence on the change. The HSI was positively associated confirmed malaria cases (OR – 1.08, Crl: 1.04, 1.12) while temperature suitability was significantly associated with confirmed cases. The small value of the precision relative to the magnitude of the cases indicates less variation in the data within the months.

There were some observed districts intervention in the nine variations in the effect of the in the region (SI Table 1). In seven out of these nine districts, the intervention was significantly associated with a reduction in malaria cases. The greatest effect was observed in Sefwi-Akontombra district where there was a 61% (OR - 0.39, CrI: 0.33, 0.47) reduction in the number of confirmed cases immediately after the intervention. Bia East, Juaboso and Bodi districts showed reductions of 49%, 38% and 32% respectively. The intervention was further associated with 27%, 22% and 17% reductions in malaria cases in Bia West, Aowin and Bibiani-Anhwiaso-Bekwai districts, respectively. However, in Sefwi-Wiawso and Suaman districts, there was no significant reduction in malaria cases due to the intervention. At district levels, habitat suitability was positively associated confirmed malaria cases in Bia East, Bibiani- Anhwiaso-Bekwai and Sefwi Akontombra districts whiles temperature suitability was negatively confirmed malaria cases in Bia East, Bia West, Bibiani-Anhwiaso-Bekwai, Sefwi Akontombra and Sefwi-Wiawso districts, respectively.

The results on the potential sustained effects of the intervention deployed after 6 months and up to 30 months is shown in Table 2. The intervention was found to have reduced malaria cases up to 30 months after the intervention. While the reduction in confirmed cases was calculated as 26% when deployed 6-months after the intervention (OR – 0.74, 95% CrI (0.65, 0.75)), there was an associated 34% (OR – 0.60 (0.49, 0.72)), 39% (OR – 0.61 (0.52, 0.71)), 37% (OR – 0.63 (0.53, 0.74)) and 40% (OR – 0.60 (0.49, 0.72)) reduction in confirmed cases at 12-, 18-, 24- and 30- months after the intervention respectively.intervention in the nine confirmed cases for the Region is depicted in Figure 4a. The plot showed a slight declining trend in monthly confirmed malaria cases before the intervention. A drastic step change in the trend post-intervention signifying reduction in the number of cases immediately after the intervention was also observed.

**Figure 4:**
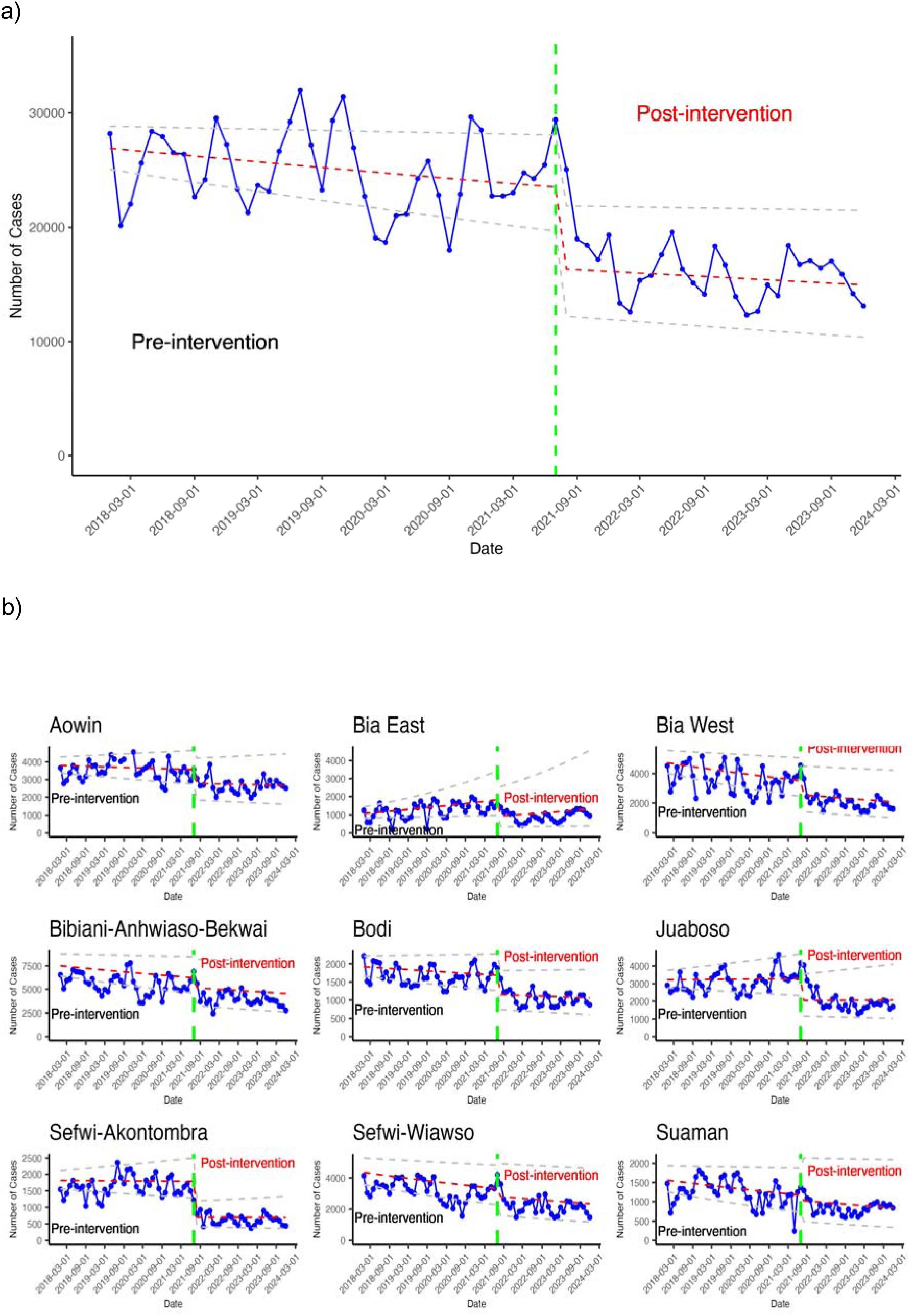
Modelled and observed time series plot of confirmed malaria cases for the regional level (a) and for districts (b) in Western North region, January 2018 to December 2023 (blue solid line with blue dots). Red dashed lines show modelled trends of cases before and after the intervention. Green vertical lines show the months where the intervention was implemented, and the grey dashed lines show the lower and upper credible intervals around the modelled estimates of the expected confirmed cases.

**Table 2:**
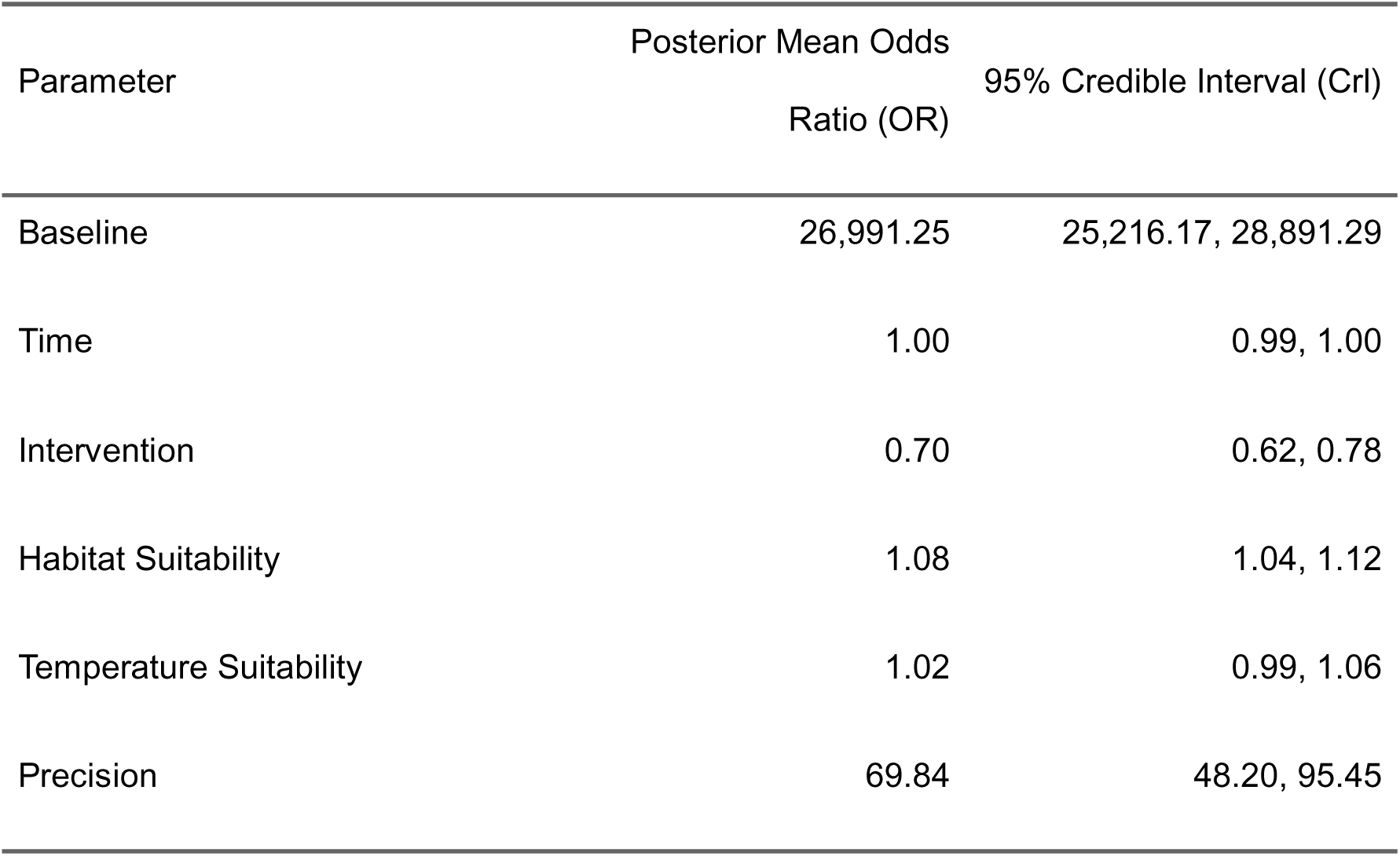
Results of Bayesian time series model of the effect of the intervention accounting for effects of exogenous variables. Baseline parameter denotes the average number of cases prior to the intervention. Time parameter is the underlining trend of cases prior to the intervention. The intervention is the time when the ITNs were deployed whiles habitat suitability and temperature suitability are the environmental covariates associated with malaria in the region. Precision values explains the level of variation in the monthly data.

**Table 3:** Results of Bayesian model analysis assessing the sustained effect of the intervention months after the distribution. Baseline parameter denotes the average number of cases prior to the intervention. Time parameter is the underlining trend of cases prior to the intervention. The parameters 6-, 12-, 18-, 24- and 30 months are six monthly time points after the intervention was deployed while precision explains the level of variation in the monthly data.

| parameter | Mean | LCI | UCI |
| --- | --- | --- | --- |
| Baseline | 25,993.35 | 24,198.14 | 27,921.72 |
| Time | 1.00 | 1.00 | 1.00 |
| 6-months | 0.74 | 0.65 | 0.84 |
| 12-months | 0.66 | 0.58 | 0.75 |
| 18-months | 0.61 | 0.52 | 0.71 |
| 24-months | 0.63 | 0.53 | 0.74 |
| 30-months | 0.60 | 0.49 | 0.72 |
| Habitat Suitability | 1.07 | 1.03 | 1.10 |
| Temperature Suitability | 1.04 | 1.00 | 1.08 |
| Precision | 75.47 | 51.42 | 104.10 |

At district levels, the intervention was found to have caused an immediate step change in trends for cases in almost all districts (Figure 4b). The step change was minimal in Sefwi-Wiawso and Suaman districts compared with the other seven districts. The plot of the modelled estimates (red dashed line) showed that cases remained relatively stable in Aowin, Bodi, Juaboso, and Sefwi-Akontombra districts post-intervention. Malaria cases, however, took a downward trend in Bia West, Bibiani-Anhwiaso-Bekwai, Sefwi-Wiawso and Suaman districts whiles an increasing trend of cases was observed in Bia East district.

## Discussion

This work explores the changes in reported confirmed malaria cases after the mass deployment of dual AI ITNs (IG2 ITNs) to households in the Western North Region of Ghana. We conducted the analysis using a Bayesian approach that allows for easy inclusion of spatial covariates and generates the levels of uncertainties around our estimates of the effect of the intervention on confirmed malaria cases in the study area. We were also able to estimate the impact the intervention had on confirmed cases up to 30 months after deployment.

We observed a slight increase in the number confirmed malaria cases between 2018 and 2019 although a mass ITN campaign was conducted in 2018, where standard pyrethroid-only nets were distributed. We however, observed a reduction of 82 537 cases between 2021 and 2022 when dual AI ITNs were distributed (Figure 1a). This suggests that dual AI ITNs had a better effect on malaria cases in the region than standard ITNs. The high stable trend in testing rates in the region indicates that confirmed cases were not affected by changes in diagnostic practices of clinicians in the region (Figure 1c). The reductions in numbers of suspected malaria cases after the intervention also suggest that malaria is a major cause of febrile illness in the region (Figure 1b). This assertion is supported by results of the 2019 MIS and the 2022 GDHS where the prevalence of fever among children less than 5 years was found to be reduced from 40.4% in the then Western region to 12.3% and 14.3% in the Western and Western North Regions, respectively(11,12). Nationally, the prevalence of fever from all causes reduced from 29.6% in 2019 to 15.1% in 2022. The declines in the number of cases in 2020 as shown in the timeseries plot (Figure 2b) and the exploratory plot (Figure 2a) can be attributed to COVID-19 pandemic disruptions in the health system. This trend was observed in most health systems across the country, as reported in previous studies(23,24). Cases surged in 2021 as COVID-19 cases declined and structures within the health system recovered from the disruptions.

We found that, the intervention reduced confirmed malaria cases by 30% immediately after deployment (Table 1). The non-significance of the trend parameter in the model suggests that the 30% reduction was associated with the intervention and was not significantly influenced by other underlying interventions in the region. ITNs use have been associated with reductions in malaria cases and deaths in most studies compared with non-use or non-treated nets(3). In Ghana, ITN usage has been found to reduce deaths from malaria in children under five years, where the under-five mortality rate was found to be 18% lower among users compared with non-users(25). The use of ITNs in areas with high insecticide resistance to pyrethroids has also been found to be associated with reduction in clinical incidence and infection prevalence(26). This assertion may however be in contrast to a study in Ghana where malaria infection prevalence was reported to be relatively higher among ITN users compared with non-users(27). Although evidence to date has not consistently demonstrated an association between level of insecticide resistance and malaria incidence or prevalence, there is credible evidence to suggest that dual AI nets have a greater killing effect on *Anopheles gambiae* than pyrethroid-only nets(28). Guided by this insight, we conclude that the observed reductions were due to the distribution of the dual AI nets, and the same effect would not have been achieved if standard pyrethroid-only nets were distributed.

The varied effectiveness of IG2 ITNs observed across the different districts likely reflects a combination of entomological, epidemiological, biochemical as well as anthropological factors. While the intervention led to significant reductions in malaria cases in seven of the nine districts, with the greatest effect observed in Sefwi- Akontombra (61% reduction), no significant impact was detected in Sefwi-Wiawso and Suaman. Differences in baseline malaria burden may explain the varying impact of dual AI ITNs across districts. Suaman, classified as a high-burden district in a 2019 stratification report, may have experienced limited impact because evidence shows that ITNs alone are often insufficient in high-transmission settings(29). Conversely, in areas with already low transmission, it may be difficult to achieve further measurable reductions. The low reductions in Sefwi-Wiawso, the most urbanised district, is postulated to have lower transmission intensity. Indeed, malaria risk is understood to be generally lower in urban areas(30,31), and this perceived risk may reduce net use as reported in other studies(32), potentially contributing to the limited impact observed. Differential coverage of PBO-deltamethrin ITNs that were co-distributed via schools, ANC, and child welfare clinics in 2020 may also have reduced chlorfenapyr efficacy in some districts as there have been reports of antagonistic effects between PBO and chlorfenapyr activation(33). Thus, in areas where PBO-LLINs are in use, there could be unintended antagonism that could reduce the effectiveness of dual AI ITNs.

Different levels of association were observed between habitat suitability and malaria cases at the regional level and in three districts, while temperature suitability was negatively associated with confirmed malaria cases in 5 districts. The reasons accounting for these phenomena is outside the objectives of this study. Results of the second model to assess the sustained effect of IG2 ITNs on malaria cases showed a consistent decline in malaria cases over the 5 periods (Table 2). Whiles confirmed malaria cases reduced by 26%, when deployed 6-months after the intervention, there was a 40% reduction in confirmed cases 30-months after the intervention. Some studies have reported a 35% reduction in malaria incidence between clusters that received dual AI ITNs and PBO and 33% reduction among clusters that received dual AI ITNs compared with their pyrethroid-only counterparts, measured 2-years post intervention(34). Typically, ITNs are intended to last three years but factors such as frequency of washing and handling can reduce their durability and potency(35). To the best of our knowledge, there is currently no published peer-review study that evaluates the effectiveness of dual AI nets on malaria risk throughout the lifespan of the nets. Nevertheless, Jacklin F. et al, 2022 reported that the insecticide effects of dual AI nets continue to be effective after 24 months, however they also found smaller reductions in malaria outcomes in the 2nd year(36). From studies suggesting the median useful life of dual AI nets to be 2.6 years, our results indicate that dual AI ITNs offer protection to the population throughout the lifespan of the net.

There are limitations to our current study and findings. Firstly, we did not account for other malaria interventions that may have been undertaken in the region unknown to the NMEP or authors although there have not been significant changes to coverages in other interventions. Health system-strengthening activities such as training and supportive supervision have been found to improve clinical practice and lead to better health outcomes(37). That said, however, such interventions could not have altered the results of our study since we accounted for underlining trends in our analysis.

Secondly, data for this study was sourced from public and private health facilities which report through the national reporting system – DHIMS2. This may not reflect the total number of malaria cases in the region as it did not include malaria services sought from unregistered public and private health facilities, pharmacies and over-the-counter-medicine sellers that do not report through the national system. We are however certain that this could not have impacted on the number of cases reported as there are few numbers of these unregistered facilities in rural areas compared with urban regions. Moreover, we did not account for changes in health-seeking behaviour over the period. From survey results in 2019 and 2022, 64% of children with fever in the past 2 weeks preceding the survey sought care in the then Western Region in 2019 while, in contrast, 71% and 51% of such did so in Western and Western North regions respectively, in 2022(11,12). It is possible that the reduction in health seeking behaviour may be attributed to the perceived risk in malaria transmission because of the intervention.

Lastly, our study did not include temporal changes in bed net use parameters or any individual level factors that influence ITN usage. The increase in ITN use in the population between 2019 and 2022 may have influenced reductions in cases as high ITN use confer herd immunity to persons who do not use nets in the same area and thereby reduces such human-vector contact. This is evidenced in a study Malawi which assessed impact of community bed net coverage and found that increased community level bed net coverage reduced malaria risk(38). Considering this evidence, such factors were not anticipated to have significantly affected the immediate impacts of the intervention as such factors may have caused gradual changes at timescales that were different to the direct effect of the ITNs.

We find that dual AI ITNs remain effective up to 30-months after distribution. Future work could extend this analysis, such as through a comparative study in similar areas that received other types of ITNs. This would improve our understanding of the effects of these nets on malaria cases under uncontrolled settings. The findings of such a future study will enhance decision making process of national malaria programs on where to deploy which type of nets. For the Ghana NMEP in particular, the findings of this work may support decisions in expanding dual AI ITN deployment to areas with high burden of malaria and confirmed pyrethroid resistance.

## Conclusion

This study demonstrated that dual AI nets were effective in reducing malaria cases in the Western North Region of Ghana under non-clinical trials settings. Dual active IG2 nets could reduce malaria cases by 40%, 30-months after distribution. As the Ghana NMEP plans to expand dual AI ITNs to other high burden areas with high insecticide resistance, we anticipate further reductions in malaria risk and success in the program to achieve its goals of reducing malaria incidence and mortality towards malaria elimination in Ghana.

### Ethics approval

The data used in this study are anonymized and aggregated, and do not include any personal identifiers. Ethical approval for this project was granted by the Ethics Committee of Curtin University (HRE2021-0734). Permission to use the data was granted by the National Malaria Elimination Programme (NMEP) of Ghana.

### Data availability

Data for this study can be provided to interested persons upon formal request to the Program Manager of the National Malaria Elimination Programme and completion of a data request form. Enquiries should be directed to

## Supplementary information

Supplementary information is available at IJE online.

## Author contributions

SO, OOA, CAO, KLM and PWG conceptualized the study. SO did the analysis and drafted the manuscript. SO, PA, KAA, PWG, reviewed the analysis and interpreted the data and results. SC, OOA, CAO, NYP contributed to the interpretation of the data and results and edited the manuscript. All authors reviewed and approved the final version of the manuscript.

## Use of Artificial intelligence (AI) tools

AI tools were used to refine R codes to add aesthetics to the plots.

## Funding

This work is part of the PhD work of the corresponding author who holds a Higher Degree by Research (HDR) full scholarship from the Curtin University.

## Acknowledgements

We are grateful to the leadership and staff of the Western North regional health directorate for implementing the net distribution. We also want to thank the staff of the National malaria elimination program who supported in the data collation and staff at the Malaria Atlas Project who helped with the covariate processing. We thank Julian Heng (ORCID: 0000-0002-0378-7078) for English-language editing of the manuscript.

## Conflict of interest

None declared

## Notes

### Competing Interest Statement

The authors have declared no competing interest.

### Funding Statement

This study did not receive any funding

### Author Declarations

Ethical approval for this project was granted by the Ethics Committee of Curtin University (HRE2021-0734). Permission to use the data was granted by the National Malaria Elimination Programme (NMEP) of Ghana

