## Supplementary information for "Reductions in malaria cases after deployment of dual-active ingredient insecticide treated nets (ITNs) in Ghana – a Bayesian interrupted time series analysis"

Supplementary information (SI)

Methods

Study data – Intervention

The Ministry of Health of Ghana uses several distribution channels to deliver ITNs to households as part of its malaria prevention efforts. These channels include routine distribution to pregnant women and children under 2 years through antenatal clinics (ANC) and child welfare clinics (CWC) in both public and private health facilities. Additionally, ITNs are distributed annually to primary school children in public and private schools - except during mass campaign years. There have been two school distribution of ITNs between 2018 and 2021. Since 2019 only PBO nets are distributed through school-based channels. Mass ITN campaigns, every three years has been the key channel of getting ITNs to all households in Ghana. During mass distribution years, community volunteers are mobilized to conduct house-to-house registration of households. The registration process follows the universal coverage concept, allocating one ITN for every two person in a household(11). Each household is given a unique identification number after registration. The registration data is collated at the district level and aggregated at regional level to determine the number of nets needed for each region. The required quantities of nets are then moved from the central level to prepositioning sites within each district. Distribution of nets to registered households occur simultaneously in all communities in a particular region. During the distribution period, registered household members go to a fixed point within the community to redeem the number of nets allocated during registration, using the unique identification number. After the distribution period, the total number of nets redeemed are collated from each community and aggregated to the regional level. In 2018, pyrethroid-only ITNs were distributed while a multi-product distribution was conducted in 2021. Three types of nets; pyrethroid-only, PBO and dual active IG2 nets, were distributed in different parts of the country guided by the level of malaria endemicity.

Study data - cases

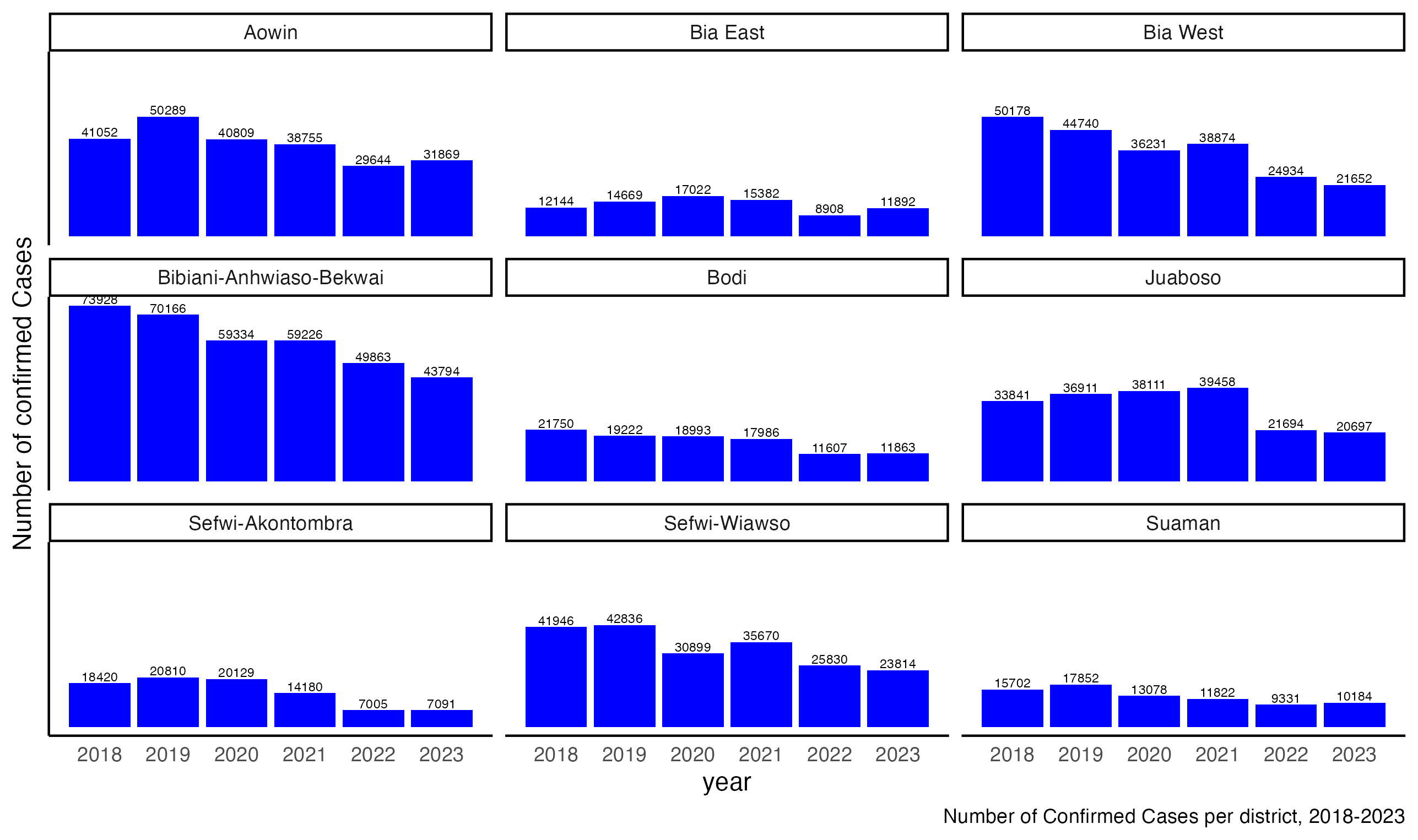

Figure 1: Yearly number of confirmed malaria cases for all nine districts in the Western North Region

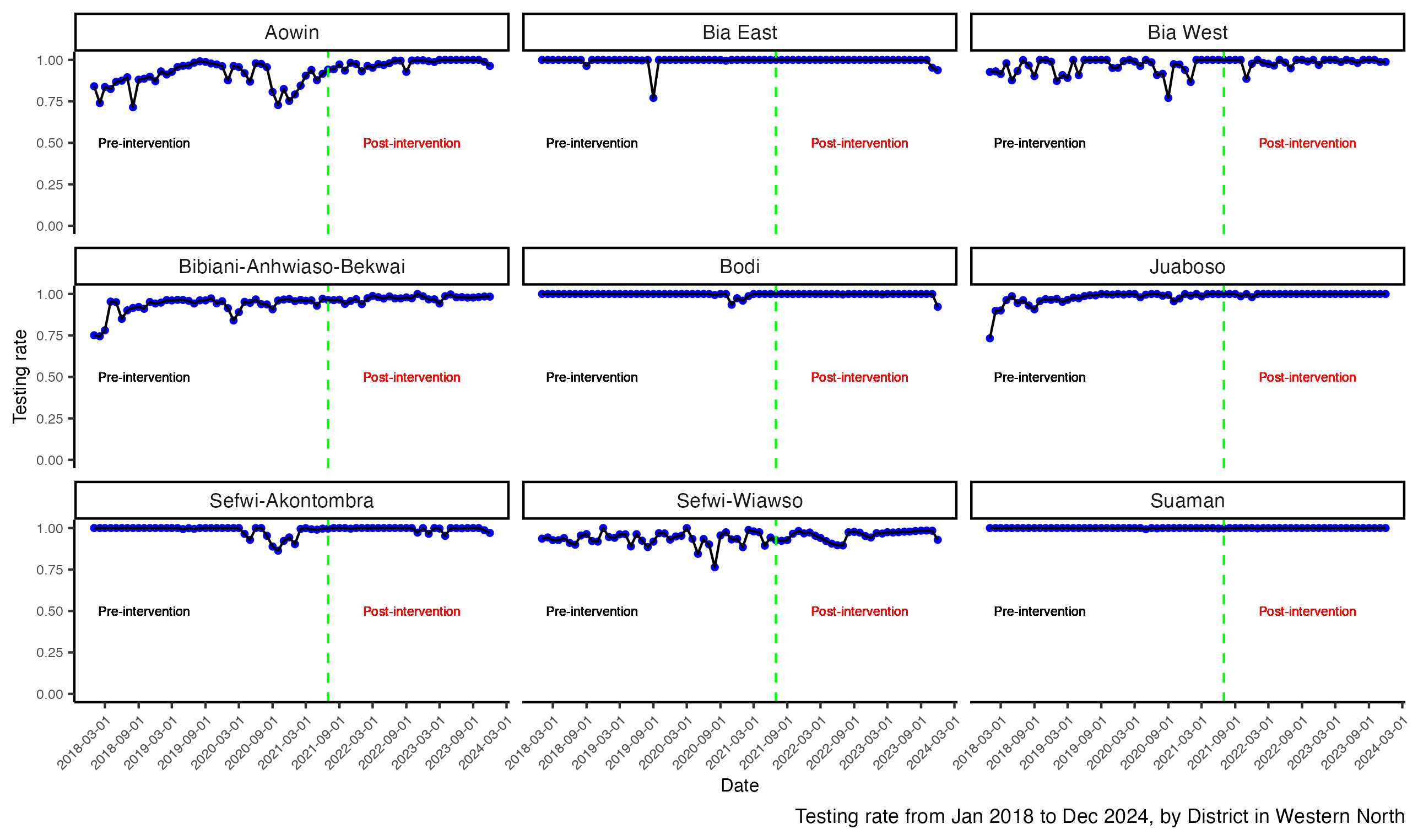

Figure 2: Trend of monthly testing rates per district pre and post intervention (black line with blue dots). Green vertical dashed line depicts month of intervention deployment.

Study data - Climatic covariates

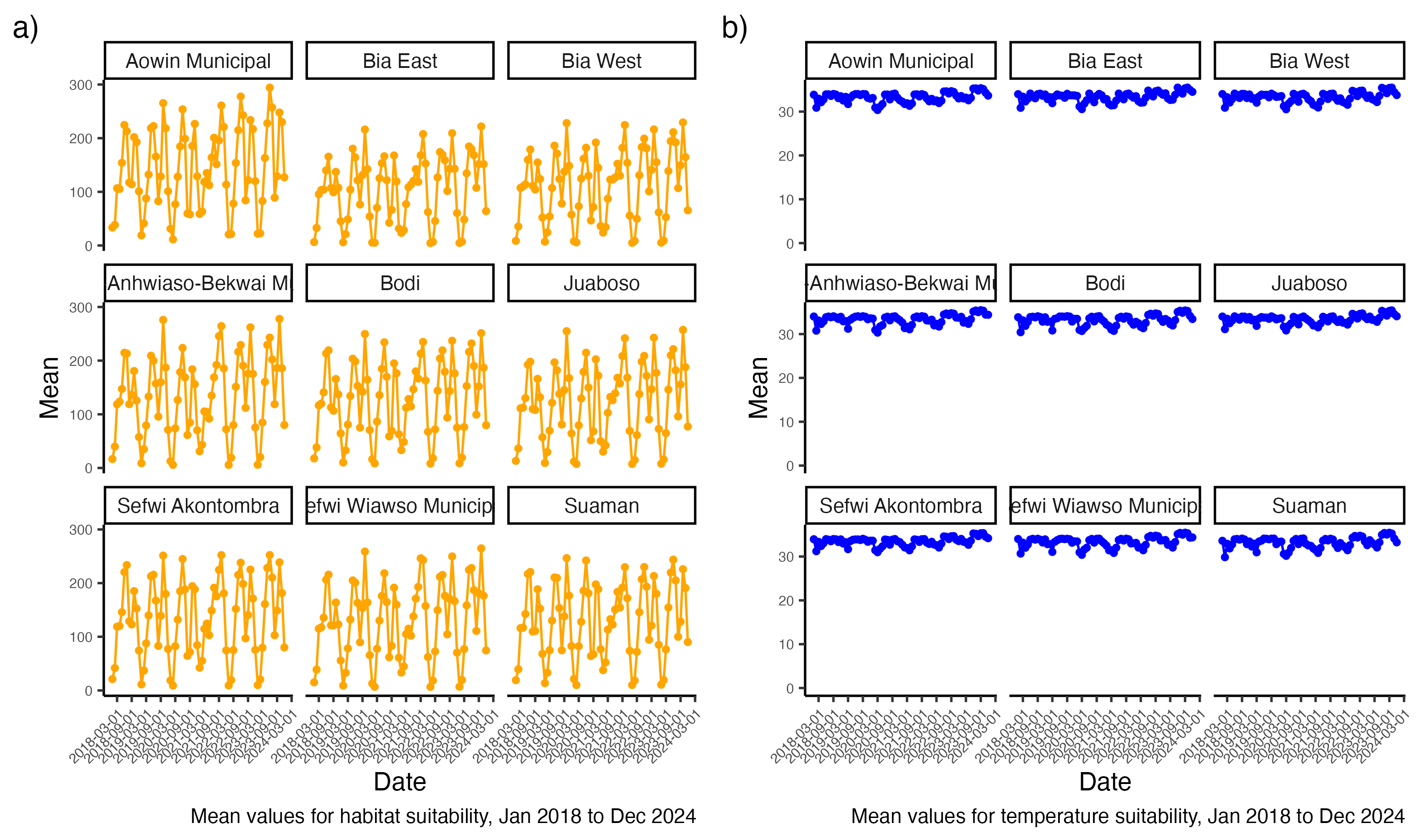

Figure 3: Mean monthly values for habitat suitability (orange) and temperature suitability by districts from January 2018 to December 2023

Bayesian Time series model

We used a Bayesian interrupted time series (BITS) model to assess the impact of the IG2 ITN distribution on malaria cases in the Western North Region. Compared to other models like auto-regressive integrated moving average (ARIMA) or interrupted time series with segmented regression (ITS-SR), BITS models are better able to handle temporal dependences in the data such as trends, seasonality and other autocorrelation effects(25). BITS allows for the inclusion of prior knowledge, better quantifies uncertainties in effect estimates and offers a more flexible approach to include exogenous variables that remain unaffected by the intervention(25,26). For this analysis we focused on what is commonly referred to as step-change also known as the level shift, and ramp(27). The step-change is a sudden change (often sustained) where the time series shifts by a given value immediately after the intervention. In our study, the step-change is denoted by the treatment variable which is coded as 0 prior to the intervention month and 1 afterwards.

$$treatment={\{}_{1, if t\geq T_{0}}^{0, if t<T_{0}}$$

Where *t* is time (months) and *T_0_* is time (month) of the intervention. The ramp, also known as the sustained effect, defines the change in slope after the intervention. In this analysis the ramp is defined as the time-since-treatment, a multifactor variable with 5 levels representing 6-monthly time points after the intervention. We built two models to explore changes in these two parameters in our dataset. The first model was to assess the step-change by the intervention while accounting for the exogenous variables namely, HSI and TSI. The second model was to determine the sustained effect of the intervention at 6 monthly intervals after the intervention.

In the first model, we assumed that the number of confirmed cases, $Y_{t}$, was Poisson distributed, given by

$Y_{t} \sim Poisson\left( \lambda_{t} \right)$

where $⋋_{t}$ is the expected confirmed cases (rate) at month *t*. We used a log-link function to ensure that the rate parameter $\lambda_{t}$ was positive. Thus, the equation for our model is given by:

$E\left( Y_{t} \right)= \lambda_{t}$,

$$g\left( \lambda_{t} \right)=log(\lambda_{t})$$

$$\log\left( \lambda_{t} \right)=\beta_{0}+ \beta_{1}T_{t}+ \beta_{2}X+ \beta_{3}HSI+ \beta_{4}TSI+ \varepsilon_{t}$$

where $\beta_{0}$ is the intercept – the baseline confirmed cases before the intervention, $\beta_{1}$ is the coefficient for the time trend $T$at month $t$, $\beta_{2}$ is the coefficient for the level change due to the intervention, $X$ is a binary indicator for the intervention which is coded 0 before the intervention and 1 after the intervention, $\beta_{3}$ is the coefficient for habitat suitability index, $\beta_{4}$ is the coefficient for temperature suitability index and $\varepsilon_{t}$ is an error term capturing residual variability.

In the second model, we modified the first model by removing the $X$ term and replacing it with the slope change term – time-since-treatment. The formula for the second model is given as

$$\log\left( ⋋_{t} \right)=\beta_{0}+ \beta_{1}* T_{t}+ \beta_{2}*Timesincetreatment{}_{1}+ \beta_{3}*Timesincetreatment{}_{2}+ \beta_{4}*Timesincetreatment{}_{3}+ \beta_{5}*Timesincetreatment{}_{4}+ \beta_{6}*Timesincetreatment{}_{5}+ \beta_{7}* HSI+ \beta_{8}* TSI+ \varepsilon_{t}$$

where $\beta_{2}, \beta_{3}, \beta_{4}, \beta_{5}, \beta_{6}$ are the coefficients for the slope change variable which is a multifactor variable with 5-levels representing 6-months, 12-months, 18-months, 24-months, and 30-months.

Model formula

The formula for the Bayesian time series model is given as;

$$its.formula=as.formula(paste\left( y \sim time+treatment+hsi+tsi \right)+$$

$$f(id, model=\text{iid}, hyper=hyper.pc))$$

where $hyper.pc$ is the list priors for the hyperparameters given as $hyper.pc=list(prior=^{'}pc.prec^{'}, params=c\left( pc.u, pc.alpha \right)))$,

$pc.u is the upper limit of the precision parameter given a value of 1$ and $pc.alpha is the probability of exceeding the upper limit given a value of 0.01$. Thus, we set a prior such that there is a probability of the parameter to exceed the value of $pc.u$.

Results

Model outputs for all districts

| Aowin | Bia East | Bia West |
| --- | --- | --- |
| parameter \| mean \| LCrI \| UCrI; Baseline \| 3,804.95 \| 3,390.16 \| 4,270.37; Time \| 1.00 \| 1.00 \| 1.00; Intervention \| 0.78 \| 0.67 \| 0.90; Habitat \| 1.04 \| 1.00 \| 1.08; Temperature \| 0.87 \| 0.76 \| 1.00; Random field \| 39.71 \| 27.34 \| 54.40 | parameter \| mean \| LCrI \| UCrI; Baseline \| 1,075.39 \| 821.81 \| 1,407.10; Time \| 1.01 \| 1.00 \| 1.02; Intervention \| 0.51 \| 0.36 \| 0.73; Habitat \| 1.20 \| 1.02 \| 1.42; Temperature \| 0.67 \| 0.52 \| 0.86; Random field \| 6.59 \| 4.55 \| 9.01 | parameter \| mean \| LCrI \| UCrI; Baseline \| 4,760.67 \| 4,067.38 \| 5,572.04; Time \| 0.99 \| 0.99 \| 1.00; Intervention \| 0.73 \| 0.59 \| 0.89; Habitat \| 1.06 \| 0.98 \| 1.16; Temperature \| 0.77 \| 0.65 \| 0.92; Random field \| 20.30 \| 14.02 \| 27.75 |
| Bibiani-Anhwiaso-Bekwai | Bodi | Juaboso |
| parameter \| mean \| LCrI \| UCrI; Baseline \| 7,536.55 \| 6,492.69 \| 8,748.31; Time \| 1.00 \| 0.99 \| 1.00; Intervention \| 0.83 \| 0.71 \| 0.97; Habitat \| 1.10 \| 1.02 \| 1.19; Temperature \| 0.65 \| 0.52 \| 0.82; Random field \| 35.10 \| 24.22 \| 48.02 | parameter \| mean \| LCrI \| UCrI; Baseline \| 1,920.89 \| 1,673.29 \| 2,205.00; Time \| 1.00 \| 0.99 \| 1.00; Intervention \| 0.68 \| 0.59 \| 0.80; Habitat \| 1.02 \| 0.96 \| 1.08; Temperature \| 0.84 \| 0.70 \| 1.02; Random field \| 36.17 \| 24.78 \| 49.82 | parameter \| mean \| LCrI \| UCrI; Baseline \| 3,216.35 \| 2,766.83 \| 3,738.85; Time \| 1.00 \| 1.00 \| 1.01; Intervention \| 0.62 \| 0.51 \| 0.76; Habitat \| 1.03 \| 0.94 \| 1.13; Temperature \| 0.88 \| 0.73 \| 1.05; Random field \| 21.61 \| 14.92 \| 29.54 |
| Sefwi-Akontombra | Sefwi-Wiawso | Suaman |
| parameter \| mean \| LCrI \| UCrI; Baseline \| 1,813.91 \| 1,564.76 \| 2,102.91; Time \| 1.00 \| 1.00 \| 1.00; Intervention \| 0.39 \| 0.33 \| 0.47; Habitat \| 1.11 \| 1.01 \| 1.21; Temperature \| 0.72 \| 0.58 \| 0.88; Random field \| 24.48 \| 16.78 \| 33.69 | parameter \| mean \| LCrI \| UCrI; Baseline \| 4,376.20 \| 3,611.33 \| 5,303.06; Time \| 0.99 \| 0.99 \| 1.00; Intervention \| 0.85 \| 0.71 \| 1.02; Habitat \| 1.09 \| 0.99 \| 1.19; Temperature \| 0.69 \| 0.51 \| 0.93; Random field \| 25.29 \| 17.44 \| 34.61 | parameter \| mean \| LCrI \| UCrI; Baseline \| 1,570.55 \| 1,273.62 \| 1,936.83; Time \| 0.99 \| 0.99 \| 1.00; Intervention \| 0.88 \| 0.68 \| 1.14; Habitat \| 0.99 \| 0.91 \| 1.07; Temperature \| 0.83 \| 0.64 \| 1.07; Random field \| 12.63 \| 8.68 \| 17.35 |
